# Modelling approaches for estimating vaccine effectiveness of consecutive SARS-CoV-2 variant sublineages in the absence of study-specific genetic sequencing data, VEBIS hospital network, Europe, 2023/24

**DOI:** 10.1101/2025.06.24.25330183

**Authors:** L Antunes, B Nunes, O Núñez, I Martínez-Baz, Y Dockx, M-L Borg, B Oroszi, R Duffy, R Duerrwald, M Kuliese, A Machado, G Petrović, M Lazăr, R Guiomar, V Álvarez Río, J Castilla, K Magerman, A Dziugyte, G Túri, M Fitzgerald, C Hackmann, L Jančorienė, V Gomez, Lovrić Makarić, O Popovici, MY Rojas-Castro, AMC Rose, the European Hospital Vaccine Effectiveness Group

## Abstract

**Introduction:** Genetic changes in COVID-19 variants/sublineages (VSLs) can reduce vaccine effectiveness (VE). Timely VSL-specific VE estimates are essential, but study-specific VSL identification by whole genome sequencing (the “gold standard”) is expensive and time-consuming. Alternatively, VSL-specific VE has been estimated from external sequencing data (VSL predominance period by proxy: PP). We propose two novel approaches for use in test-negative design (TND) studies to estimate VSL-specific VE when study-specific VSL identification is not possible.

**Methods:** We demonstrate the variant category model (VCM) and the variant proportion model (VPM) approaches. Using data from a hospital-based TND study among adults ≥65 years, during the period of sequential predominance of XBB and BA.2.86 in 2023/24, we estimated the VE as (1-OR) x 100%. For the VCM, we used a binary variable categorising “most likely underlying sublineage” based on publicly available sequencing data. For the VPM, we used a continuous variable with values from 0 to 1 representing the weekly proportion of BA.2.86. We validated results using study-specific VSL identification from sequenced study data (SD) and the standard PP approach.

**Results:** Overall, at 14–59 days post vaccination, VE point estimates against XBB were within ±3% absolute for the VE estimated using both models, with an equivalent standard PP validation. We could not validate using SD, as there were no vaccinated XBB cases. Against BA.2.86, VE was lower than against XBB, and the VCM and VPM results were within ±7% absolute of each other, with lowest validation results from SD but equivalent results from the PP.

**Conclusions:** Both proposed approaches produced similar VE estimates to those from well-known methods. The VPM could also provide VE estimates when the validation techniques were limited by low sample size.

## Introduction

Genetic changes in SARS-CoV-2 have been shown to affect viral characteristics such as transmissibility, virulence, and immune evasion, as well as therapeutic response of emerging SARS-CoV-2 variants [1,2]. The emergence of new variants/sublineages (VSLs) may result in reduced vaccine effectiveness (VE), increased risk of disease severity (particularly among older and other at-risk populations [1]), and compromised healthcare systems. VSL-specific VE estimation is key for the development of vaccines to ensure continued protection as the virus evolves, particularly among those most vulnerable. Timely VSL-specific VE estimates may inform immunisation programmes to be adapted and mitigate the impact of the newly emerging variants.

There are three ways to monitor existing, and identify emerging variants, of which the gold standard is whole genome sequencing [3]. Methods to estimate VE against a VSL of interest previously relied on study-specific VSL identification using either sequencing data from a specimen collected from study participants, linked with epidemiological data [4,5] or inferred from RT-PCR techniques [6,7]. Although most VE studies require PCR confirmation of cases, VSL identification is not a requirement for patient treatment. Adding a sequencing component to VE studies requires additional efforts in terms of logistics and cost, and can impact the timeliness of results [4]. In addition, linkage of general surveillance sequencing to study data, particularly for studies collecting primary data, can be difficult. Sequencing data may not be available when VE estimates are prepared and, even when available, if there is only a low number of positive samples sequenced, this can limit precision of the VSL-specific VE estimates [4]. During the COVID-19 pandemic period, some studies began using a proxy to genetically classify viruses and VSLs, by restricting the study period to when the virus or VSL of interest was predominant (often with varying predominance thresholds), based on external data sources [8–13].

During the 2023/24 COVID-19 vaccination campaign in the European Union/European Economic Area (EU/EEA), newly adapted monovalent XBB.1.5 COVID-19 vaccines (XBB.1.5 vaccines) accounted for the majority of all known vaccines administered [14]. The SARS-CoV-2 XBB VSLs had been dominant in Europe from before the start of the vaccination campaign in 2023 until mid-December, when BA.2.86 VSLs (in particular JN.1) became predominant [15]. Since BA.2.86 VSLs were associated with potential immune escape [16], estimating the effectiveness of the XBB.1.5 vaccine against this new SARS-CoV-2 VSL was of utmost importance, together with assessing any difference in VE against XBB vs the subsequently predominant BA.2.86 VSLs. Some studies estimated VE against these time-consecutive SARS-CoV-2 VSLs separately using sequenced data, and compared estimates [4,5], while others directly compared the odds of vaccination between cases infected with the different VSLs, in case–case or case-only studies [17,18].

Here we propose two novel modelling approaches for estimating VE against time-consecutive VSLs (i.e. when there is a transition from one predominant VSL to another) when study-specific sequenced data is unavailable and proxy external data sources are used to identify predominantly circulating VSLs. To illustrate this, we used data from a European test-negative design study conducted in 2023/24 [11,19]. The proposed models permit a statistical comparison of the VE against two consecutive VSLs as well as VE estimation of each. In order to validate our proposed models, we compare the modelling results with those from the gold standard approach based on VSL cases classified using sequencing, and against the VE for each predominant period by using external data as a proxy.

## Methods

### Study design

The proposed modelling approaches were developed and applied in the context of test-negative design studies in the hospital setting to estimate VE. Very briefly, in this study design and setting [20], hospitalised patients are systematically recruited based on a clinical definition of severe acute respiratory infection (SARI) and undergo a laboratory test (preferably RT-PCR) for the target pathogen. Patients with an RT-PCR-positive result for SARS-CoV-2 are classified as “cases” and patients with a negative result as “controls”. Vaccine effectiveness is estimated as VE=(1-OR) x 100%, where OR is the estimated odds ratio of vaccination between cases and controls, adjusted for confounding, and obtained using logistic regression models.

### Usual approaches for VSL-specific VE estimation

We consider the gold standard approach to estimate the VSL-specific VE as when VSL cases within the study are classified using sequenced data (“SD method”). We consider the VSL predominance period approach (“standard PP”) when VSL-specific VE is estimated using an external sequenced data source to classify VSL cases. In this approach, the study period is restricted to the period of predominance of the VSL of interest, i.e. all cases are considered to have been likely infected by a VSL of interest if the date of onset/swab falls within the period of that VSL predominance. Usually, the VSL predominance period is defined as the sequential weeks where the proportion of all sequenced viruses belonging to the VSL under study is above a predefined threshold (e.g. 60%), calculated using population-level surveillance data from an external, publicly available source (e.g. GISAID [21], European Respiratory Virus Surveillance Summary (ERVISS) [22]). For both of these methods, VE is calculated in the standard way as described in the previous section.

### New modelling approaches

Our proposed novel modelling approaches for estimating VSL-specific VE use an “adapted PP” method, during the period of transition from the predominance of one VSL to another. For this purpose, instead of restricting the analysis period to each individual VSL predominance period determined from an external source and performing an analysis for each period, we introduce a new independent variable in the regression model to represent the VSL. This can be done in two ways: (1) as a categorical variable, that assumes the value of 0 for participants recruited during the period of predominance of the earlier VSL, and 1 for participants recruited during the period of predominance of the subsequent VSL (the “VSL category model”: VCM); and (2) as a continuous variable with values ranging from 0 to 1, representing the proportion of the subsequent VSL (the VSL gaining dominance) out of all sequenced viruses each week (the “VSL proportion model: VPM”). We summarise the four methods/approaches in Table 1.

**Table 1.** General description of the usual standard techniques and proposed novel model techniques to estimate variant/sublineage (VSL)-specific VE, VEBIS hospital study, Europe.

| Method/ model | Description |
| --- | --- |
| **Gold standard SD** (sequenced data) | Sequenced data approach: the gold standard, using actual sequencing results linked to epidemiological results in the dataset |
| **Standard PP** (standard predominance period) | Predominance period approach: standard technique using a proxy for the linked sequenced data, based on external VSL sequenced data sources where cutoff dates for variant/sublineage (VSL) predominance are determined and used to restrict the analysis period to the VSL of interest |
| **VCM** (VSL category model) | First novel approach: VSL category model, also based on external VSL sequenced data sources, in which a binary variable is used to define the predominant VSL periods (taking a value of 0 during the first VSL predominance period and 1 for the second) |
| **VPM** (VSL proportion model) | Second novel approach: VSL proportion model, also based on external VSL sequenced data sources, in which a continuous variable is used to define the proportion of the VSL of interest, or the VSL gaining predominance (ranging from 0 to 1, where 0 and 1 represent respectively the absence and total dominance of the new emerging VSL) |

VEBIS: Vaccine Effectiveness, Burden and Impact Studies; VSL: variant/sublineage.

### Test scenario

To exemplify the described approaches to estimate VSL-specific VE, we used data from patients hospitalised with SARI from the Vaccine Effectiveness, Burden and Impact Studies (VEBIS) hospital network from September 2023 to April 2024, covering the period of transition between SARS-CoV-2 VSLs XBB to BA.2.86 predominance. Details of the methods used for standard COVID-19 VE analyses in the VEBIS hospital study are already published [11,19,23] as well as the results from this period [19].

Definitions used in current analyses (including of vaccinated and unvaccinated patients, and analytic exclusions), as well as a description of the proxy determination for the XBB and BA.2.86 predominance periods, can be found in Supplementary Table S1 and Section S1, respectively. We defined each study site-specific start study period as 14 days after the introduction of the XBB.1.5 vaccine within each country (Supplementary Table S2); there were also further restrictions depending on the modelling approach (see descriptions of each approach below). We estimated VE against each of the XBB and BA.2.86 VSLs as a “test scenario” using the VCM and the VPM modelling techniques. For “BA.2.86” we include all of its sublineages, such as JN.1. We validated our results by also estimating VE using both SD and the standard PP methods (when sample size allowed; see next section).

We restricted analysis to those vaccinated <60 days before symptom onset (where the period of the VSLs overlap) and to patients aged ≥65 years.

### Statistical analysis

For all approaches (SD, standard PP, VCM, and VPM), we first estimated the OR of vaccination between cases and controls with study site as a fixed effect and adjusted for confounding by date of symptom onset, sex, age and presence of at least one chronic condition (diabetes, heart disease, lung disease/asthma and immunodeficiency). The best functional forms of the continuous variables age and onset date (categories, splines, linear terms) were selected using the Akaike information criterion (AIC). We estimated VE overall (14–59 days) and by time since vaccination (TSV) in 30-day bands (14–29 and 30–59 days post vaccination). For the TSV analysis, vaccination was included as a categorical variable (unvaccinated, 14–29 days, 30–59 days post vaccination).

As described previously [11], we do not show estimates if there were no vaccinated cases, <20 vaccinated patients or when the VE estimate had an absolute difference >10% from the VE estimated using penalised logistic regression (to assess small sample bias). We carried out a complete case analysis.

### The SD (sequenced data) approach

Here, cases were defined as those with a sequenced biological sample and classified as SARS-CoV-2 VSLs BA.2.86 or XBB. As the XBB period was earlier, to estimate XBB.1.5 VE against XBB VSL, we restricted the end of this study period for analysis as the date of onset of the last sequenced XBB case, at study site level. To estimate XBB.1.5 VE against BA.2.86 VSL, we defined the start of the study period as the date of onset of the first BA.2.86 sequenced case and the end as the date of onset of the last patient included at the time of the analysis. Since each study period was defined by the duration of the identification of the VSL of interest by sequencing, there could be an overlap in study periods and consequently, in each study site, some controls included in the models for XBB might also be included for BA.2.86.

### PP (standard VSL predominance period) approach

In the standard PP approach, we obtained XBB.1.5 VE against XBB and BA.2.86 VSLs by restricting the analysis to the periods of XBB and BA.2.86 predominance, respectively. Two models were fitted to the data restricted to each VSL predominance period to estimate each VSL VE. The definition of the study periods for XBB and BA.2.85 predominance is described in detail in Supplementary Section S1. Briefly, we used a threshold of 60% samples sequenced (using publicly available data from ERVISS 21]) to determine the weeks in each country during which each VSL was predominant (shown visually in Figure 1 (a) and (b), and with start dates listed in Supplementary Table S3).

**Figure 1.**
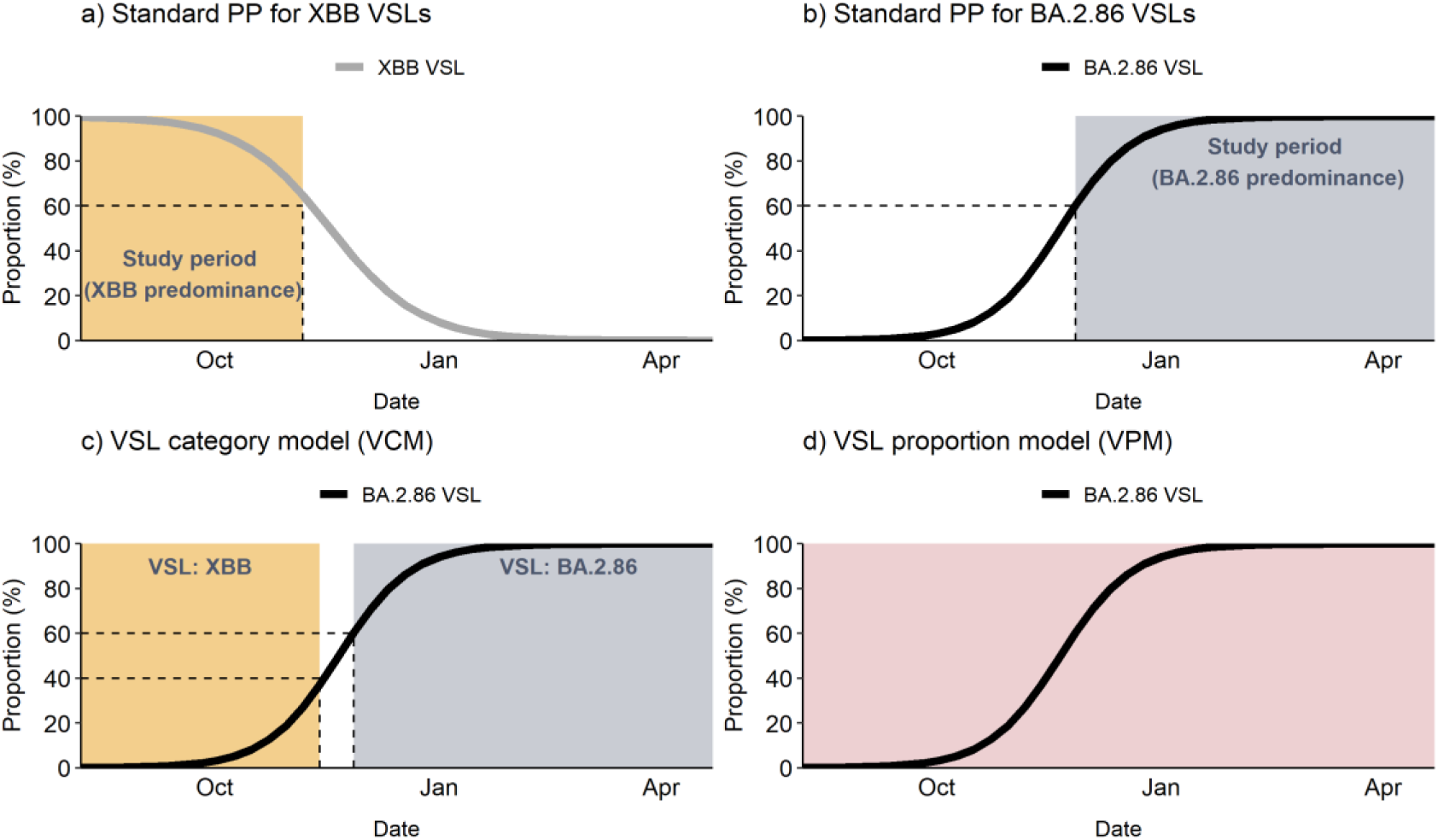
Demonstration of the study period definitions for the predominance period approaches: standard PP VE against XBB VSL (a), standard PP VE against BA.2.86 (b) and novel model approaches: the VSL category (0 if XBB and 1 if BA.2.86) model (c) and VSL proportion (prevalence of BA.2.86 VSL) model (d). VEBIS hospital network, Europe Standard PP: predominance period approach – standard technique using proxy for sequenced data; VCM: novel VSL category model; VEBIS: Vaccine Effectiveness, Burden and Impact Studies; VPM: novel VSL proportion model; VSL: variant/sublineage.

### Adapted PP VCM (predominance period with VSL categorical variable) approach

In the proposed adapted PP with VCM, we included in the model a binary “VSL variable” to represent the most likely underlying VSL causing the infection (0 for XBB, 1 for BA.2.86 in this test scenario), and the interaction term between the VSL and vaccination variables (Eq. 1).

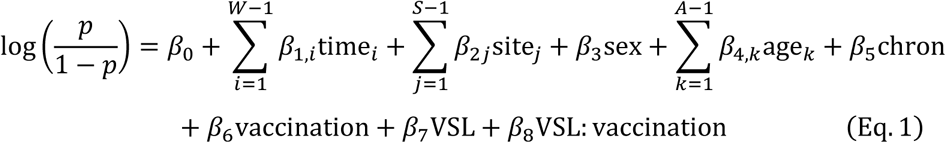

In this test scenario, the VSL variable represents XBB and BA.2.86 when the weekly BA.2.86 VSL proportion is ≤40% or ≥60%, respectively (Figure 1 (c)). *W* represents the number of weeks of the study period, *S*, the number of sites and *A* the number of age groups, assuming time and age as discrete variables (the best functional form of these variables based on the AIC might differ across models).

Based on model Eq. 1 and for this test scenario, VE against the XBB VSL is calculated as (1 − *OR_XBB_*) × 100%, where the OR of vaccination against XBB (*OR_XBB_*) is given by *e^β^*_6_. Similarly, the VE against the BA.2.86 VSL can be calculated as (1 − *OR*_*BA*.2.86_) × 100%, where the OR of vaccination against BA.2.86 (*OR*_*B.A*.2.86_) is given by 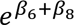.

To statistically compare the VE against each VSL, we can calculate the ratio of the ORs (ROR), 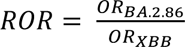, which is given by the exponential of the interaction term in the model (*e*^*β*^_8_), and its *p*-value. The ROR is a relative measure used to compare the OR from two groups, i.e., to assess if the relationship between an exposure of interest and an outcome differs between two groups. If the *ROR* = 1 , then there are no differences in VE against XBB and BA.2.86 VSLs; if *ROR* > 1, the OR against BA.2.86 is higher than the OR against XBB VSLs (so the VE against BA.2.86 is lower than against XBB VSLs); and if *ROR* < 1, the VE against BA.2.86 is higher than against XBB VSLs.

It can be also proven that the OR of vaccination estimated by case–case modelling approaches [24], when controls are the same, corresponds to the ratio of the OR between the two VSLs 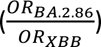 (Supplementary Table S4).

For the models used to estimate VSL VE by TSV, deductions can be made in a similar way, noting that now that the vaccination variable has more than two categories.

### Adapted PP VPM (VSL as a continuous variable) approach

In the proposed adapted PP with VPM approach, the VSL variable in Eq. 1 is represented as a continuous numeric variable with values ranging from zero to one, depicting the population level relative frequency of SARS-CoV-2 cases that belong to the VSL BA.2.86. This modelling approach allows the estimation of the VSL-specific VE at any value of the BA.2.86 VSL proportion (Figure 1 (d)). In Eq. 1, if we assume VSL as the proportion of cases classified as BA.2.86 VSL, the OR of being vaccinated in cases vs controls can be written as a function of the BA.2.86 VSL proportion of interest *x*, as presented below:

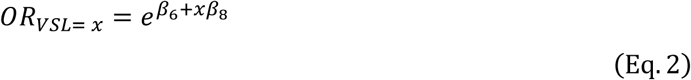

When *x* = 0, VE is being estimated during weeks when the prevalence of BA.2.86 is null, if all the SARS-CoV-2 cases belonged to XBB VSL, 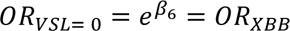. Likewise, when *x* = 1 , VE is being estimated during weeks when the proportion of BA.2.86 is 1, so 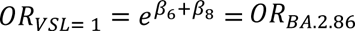.

For both the VCM and VPM approaches, the BA.2.86 VSL proportion weekly time series at study level was smoothed by fitting a logistic regression to the weekly proportion of BA.2.86 VSL extracted from ERVISS (Supplementary Figure S1). If the study site weekly BA.2.86 proportion was missing, the values were imputed using data from neighbouring countries (to overcome weeks with missing data and/or with a low number of sequenced samples).

As a sensitivity analysis, to enable comparison of results from the VPM with the other approaches, we estimated the OR when VSL = 0.3 for the VE against XBB and VSL = 0.7 for the VE against BA.2.86. More specifically 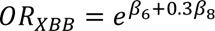 and 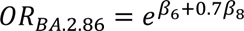.

## Results

Within our “test scenario” period (September 2023–April 2024), the start date for data included in both SD and standard PP XBB analyses was 02 October 2023, while the BA.2.86 start dates were 09 October 2023 (for SD analyses) and 22 November 2023 (for standard PP analysis). End dates were in December 2023 for XBB analyses (07 December for PP and 19 December for SD analyses) and in April 2024 for BA.2.86 (25 April for SD and 26 April for standard PP). For both VCM and VPM approaches, the study period was from 02 October 2023 to 26 April 2024. The final study periods, number of SARI patients included and number and sites included in the standard technique and novel model analyses are shown in Table 2.

**Table 2.** Final study periods, numbers of SARI patients and number and sites included in analyses using standard techniques and proposed novel model techniques, VEBIS hospital study, Europe.

| Method/<br>model | VSL | Study<br>period start | Study period<br>end | Number of<br>included SARI<br>patients | Number and<br>sites in<br>analysis | Supplementary<br>figures<sup>a</sup> |
| --- | --- | --- | --- | --- | --- | --- |
| Gold<br>standard<br>SD | XBB | 02 Oct 2023 | 19 Dec 2023 | 1011 | Two (ES, HR) | Figure S2 |
|  | BA.2.86 | 09 Oct 2023 | 25 Apr 2024 | 3486 | Five (BE, DE, ES, IE, NA) | Figure S3 |
| Standard<br>PP | XBB | 02 Oct 2023 | 07 Dec 2023 | 491 | Four (DE, ES, HR, NA) | Figure S4 |
|  | BA.2.86 | 22 Nov 2023 | 26 Apr 2024 | 3981 | Six (BE, DE, ES, IE, MT, NA) | Figure S5 |
| VCM | XBB and<br>BA.2.86 | 02 Oct 2023 | 26 Apr 2024 | 4795 | Seven (BE, DE, ES, HR, IE, MT, NA) | Figure S6 |
| VPM | XBB and<br>BA.2.86 | 02 Oct 2023 | 26 Apr 2024 | 5215 | Seven (BE, DE, ES, HR, IE, MT, NA) | Figure S7 |

SARI: severe acute respiratory infection; BE: Belgium; DE: Germany; ES: Spain; HR: Croatia; IE: Ireland; MT: Malta; NA: Navarre; PP: predominance period approach – standard technique using proxy for sequenced data; SARI: severe acute respiratory infection; SD: sequenced data (gold standard technique); VCM: novel VSL category model; VEBIS: Vaccine Effectiveness, Burden and Impact Studies; VPM: novel VSL proportion model; VSL: variant/sublineage.
<sup>a</sup> Figures in Supplementary material show the exclusion flowcharts for each technique and VSL, for each study period.

### Vaccine effectiveness results

Overall, in the first 14–59 days post vaccination, as there were no vaccinated cases in the SD analysis for VE against XBB VSLs, we could only validate the models against the standard PP (Table 3).

**Table 3.**
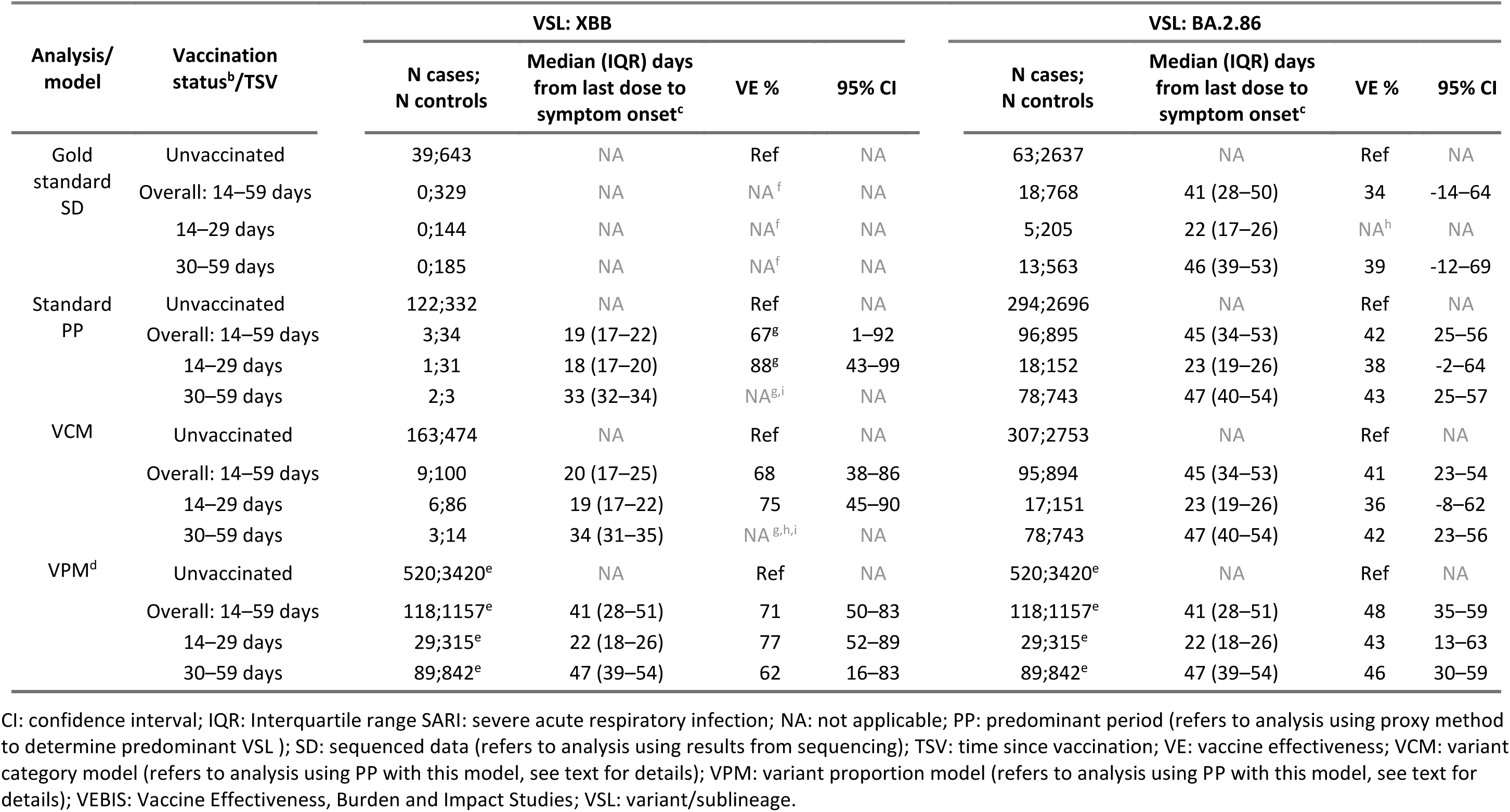
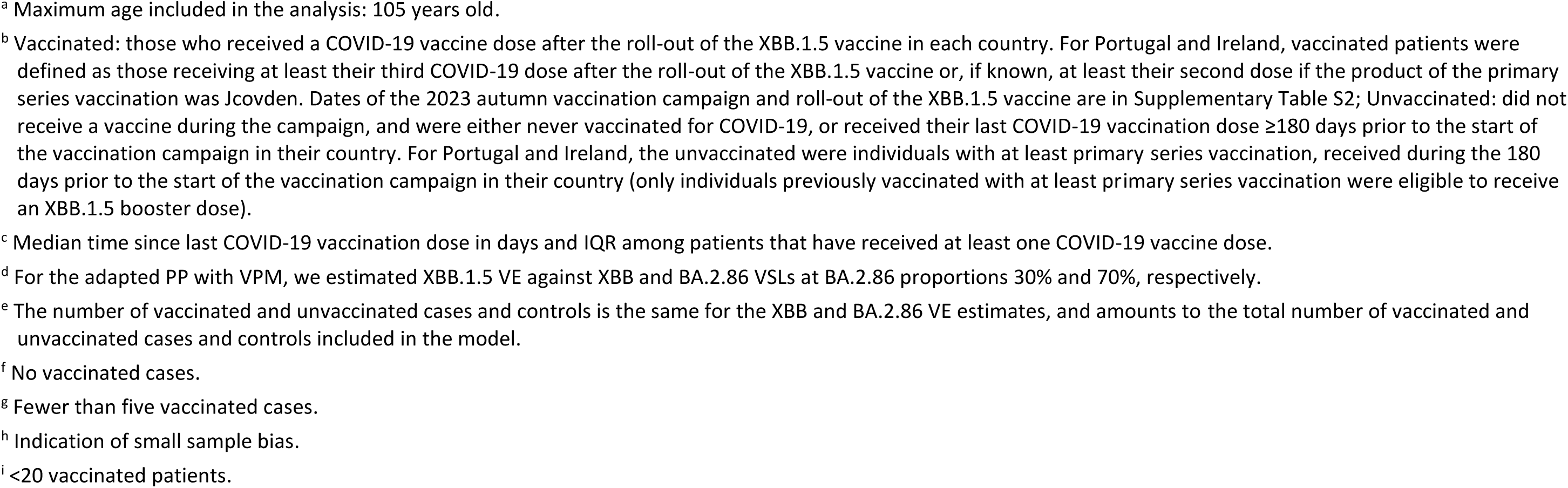
Effectiveness of the monovalent XBB.1.5 COVID-19 vaccines against hospitalisation among individuals aged ≥65 years old^a^, by VSL (XBB or BA.2.86) and by time since vaccination (TSV), using four modelling approaches: sequence data (SD), standard predominance period approach (PP), adapted predominance period with VSL categorical variable approach (VCM) and with VSL proportion variable approach (VPM). VEBIS hospital network, Europe, 2023/24

| Analysis/<br>model | Vaccination<br>status<sup>b</sup>/TSV | VSL: XBB |  |  |  | VSL: BA.2.86 |  |  |  |
| --- | --- | --- | --- | --- | --- | --- | --- | --- | --- |
|  |  | N cases;<br>N controls | Median (IQR) days<br>from last dose to<br>symptom onset<sup>c</sup> | VE % | 95% CI | N cases;<br>N controls | Median (IQR) days<br>from last dose to<br>symptom onset<sup>c</sup> | VE % | 95% CI |
| Gold<br>standard<br>SD | Unvaccinated | 39;643 | NA | Ref | NA | 63;2637 | NA | Ref | NA |
|  | Overall: 14–59 days | 0;329 | NA | NA<sup>f</sup> | NA | 18;768 | 41 (28–50) | 34 | -14–64 |
|  | 14–29 days | 0;144 | NA | NA<sup>f</sup> | NA | 5;205 | 22 (17–26) | NA<sup>h</sup> | NA |
|  | 30–59 days | 0;185 | NA | NA<sup>f</sup> | NA | 13;563 | 46 (39–53) | 39 | -12–69 |
| Standard<br>PP | Unvaccinated | 122;332 | NA | Ref | NA | 294;2696 | NA | Ref | NA |
|  | Overall: 14–59 days | 3;34 | 19 (17–22) | 67<sup>g</sup> | 1–92 | 96;895 | 45 (34–53) | 42 | 25–56 |
|  | 14–29 days | 1;31 | 18 (17–20) | 88<sup>g</sup> | 43–99 | 18;152 | 23 (19–26) | 38 | -2–64 |
|  | 30–59 days | 2;3 | 33 (32–34) | NA<sup>g,i</sup> | NA | 78;743 | 47 (40–54) | 43 | 25–57 |
| VCM | Unvaccinated | 163;474 | NA | Ref | NA | 307;2753 | NA | Ref | NA |
|  | Overall: 14–59 days | 9;100 | 20 (17–25) | 68 | 38–86 | 95;894 | 45 (34–53) | 41 | 23–54 |
|  | 14–29 days | 6;86 | 19 (17–22) | 75 | 45–90 | 17;151 | 23 (19–26) | 36 | -8–62 |
|  | 30–59 days | 3;14 | 34 (31–35) | NA<sup>g,h,i</sup> | NA | 78;743 | 47 (40–54) | 42 | 23–56 |
| VPM<sup>d</sup> | Unvaccinated | 520;3420<sup>e</sup> | NA | Ref | NA | 520;3420<sup>e</sup> | NA | Ref | NA |
|  | Overall: 14–59 days | 118;1157<sup>e</sup> | 41 (28–51) | 71 | 50–83 | 118;1157<sup>e</sup> | 41 (28–51) | 48 | 35–59 |
|  | 14–29 days | 29;315<sup>e</sup> | 22 (18–26) | 77 | 52–89 | 29;315<sup>e</sup> | 22 (18–26) | 43 | 13–63 |
|  | 30–59 days | 89;842<sup>e</sup> | 47 (39–54) | 62 | 16–83 | 89;842<sup>e</sup> | 47 (39–54) | 46 | 30–59 |

CI: confidence interval; IQR: Interquartile range SARI: severe acute respiratory infection; NA: not applicable; PP: predominant period (refers to analysis using proxy method to determine predominant VSL); SD: sequenced data (refers to analysis using results from sequencing); TSV: time since vaccination; VE: vaccine effectiveness; VCM: variant category model (refers to analysis using PP with this model, see text for details); VPM: variant proportion model (refers to analysis using PP with this model, see text for details); VEBIS: Vaccine Effectiveness, Burden and Impact Studies; VSL: variant/sublineage.
<sup>a</sup> Maximum age included in the analysis: 105 years old.
<sup>b</sup> Vaccinated: those who received a COVID-19 vaccine dose after the roll-out of the XBB.1.5 vaccine in each country. For Portugal and Ireland, vaccinated patients were defined as those receiving at least their third COVID-19 dose after the roll-out of the XBB.1.5 vaccine or, if known, at least their second dose if the product of the primary series vaccination was Jcovden. Dates of the 2023 autumn vaccination campaign and roll-out of the XBB.1.5 vaccine are in Supplementary Table S2; Unvaccinated: did not receive a vaccine during the campaign, and were either never vaccinated for COVID-19, or received their last COVID-19 vaccination dose $\geq 180$ days prior to the start of the vaccination campaign in their country. For Portugal and Ireland, the unvaccinated were individuals with at least primary series vaccination, received during the 180 days prior to the start of the vaccination campaign in their country (only individuals previously vaccinated with at least primary series vaccination were eligible to receive an XBB.1.5 booster dose).
<sup>c</sup> Median time since last COVID-19 vaccination dose in days and IQR among patients that have received at least one COVID-19 vaccine dose.
<sup>d</sup> For the adapted PP with VPM, we estimated XBB.1.5 VE against XBB and BA.2.86 VSLs at BA.2.86 proportions 30% and 70%, respectively.
<sup>e</sup> The number of vaccinated and unvaccinated cases and controls is the same for the XBB and BA.2.86 VE estimates, and amounts to the total number of vaccinated and unvaccinated cases and controls included in the model.
<sup>f</sup> No vaccinated cases.
<sup>g</sup> Fewer than five vaccinated cases.
<sup>h</sup> Indication of small sample bias.

The VE against XBB VSLs for this period was 68% (95%CI: 38–86) using the VCM, 71% (95%CI: 50–83) with the VPM and, in validation, 67% (95%CI: 1–92) using standard PP (Figure 2). Against BA.2.86 VSLs, VE was 41% (95%CI: 23–54) using the VCM, 48% (95%CI: 35–59) with the VPM and, for validation, VE was 34% (95%CI: -14–64) in the SD analysis and 42% (95%CI: 25–56) using standard PP.

**Figure 2.**
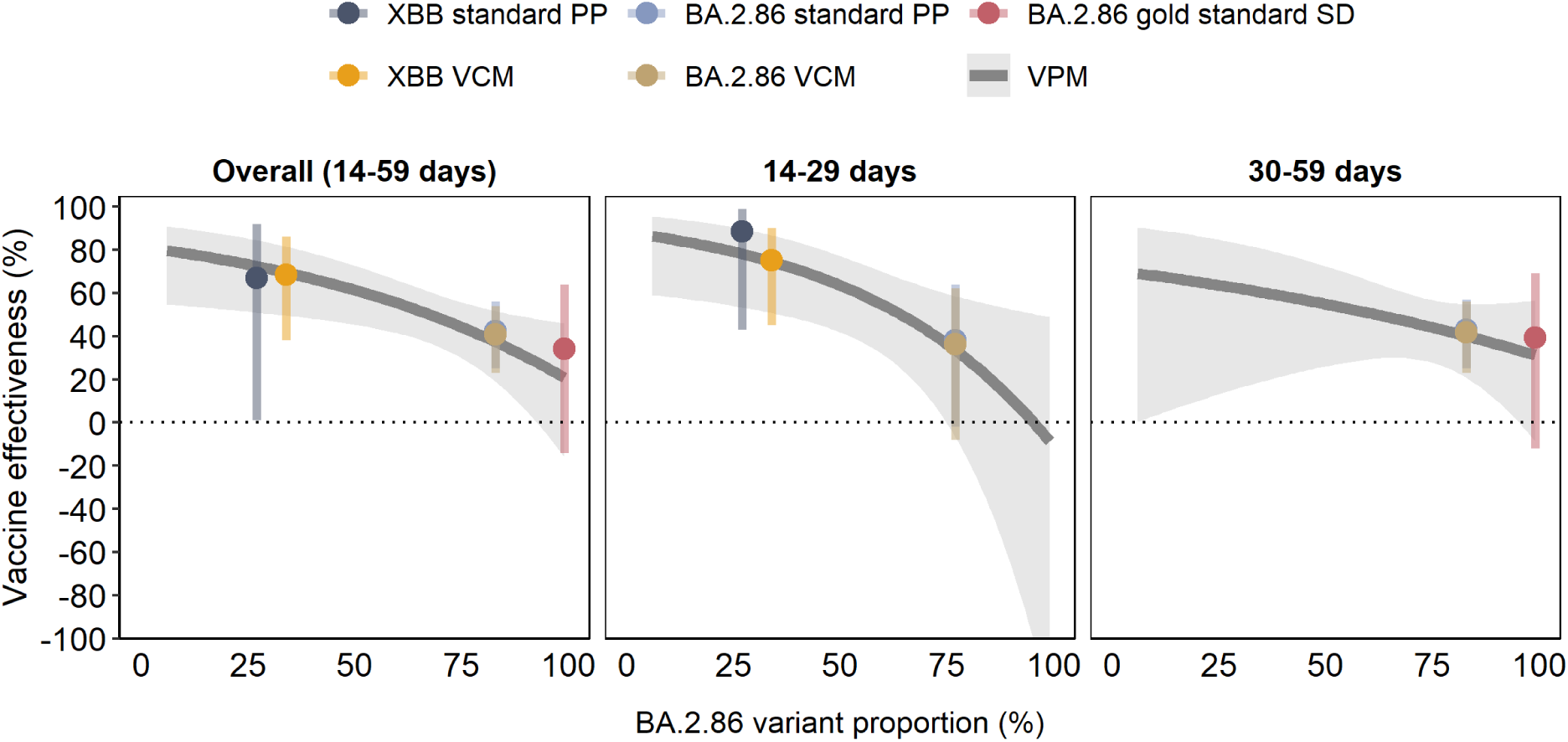
Effectiveness of the monovalent XBB.1.5 COVID-19 vaccines against hospitalisation among individuals aged ≥65 years old, by BA.2.86 VSL proportion and by time since vaccination, using the four approaches: the gold standard sequencing data (gold standard SD), standard predominance period (standard PP), adapted predominance period with VSL category model (PP VCM) and with the proportion model (PP VPM); VEBIS hospital network, Europe

At 14–29 days post vaccination, due to small sample sizes, the VE using SD was not possible for either VSL and the models could only be validated using standard PP. Against XBB VSL, VE was 75% (95%CI: 45 –90) using the VCM, and 77% (95%CI: 52 –89) using the VPM, while the standard PP validation VE was 88% (95%CI: 43 –99). Against BA.2.86 VSL, VE was 36% (95%CI: -8–62) for the VCM, 43% (95%CI: 13–63) using the VPM, and 38% (95%CI: -2–64) in validation using standard PP.

At 30–59 days post vaccination, low sample size precluded any estimates using the VCM. The VE against XBB VSLs was 62% (95%CI: 16–83) using the VPM. Against BA.2.86 VSL, VE was 42% (95%CI: 23–56) using the VCM, and 46% (95%CI: 30–59) using the VPM. Validation using SD was 39% (95%CI: -12–69) and 43% (95%CI: 25–57) using standard PP.

The results from the proposed methods, and well as from the standard techniques, show a decline in VE as BA.2.86 VSLs increased in circulation, overall and by TSV (Figure 2).

### Comparison between VE against XBB and BA.2.86 VSLs

The ROR against BA.2.86 and XBB VSLs in the first 14–59 days post vaccination was 1.9 (95%CI: 0.9– 4.3) and 1.8 (95%CI: 1.1–2.9) using the VCM and the VPM, respectively (Table 4). At 14–29 days post vaccination, the ROR against BA.2.86 vs XBB VSLs was 2.6 (95%CI: 1.0–7.6) with the VCM and 2.4 (95%CI: 1.2–5.0) with the VPM. At 30–59 days post vaccination, the ROR against BA.2.86 vs XBB VSLs was 1.4 (95%CI: 0.7–2.7) using the VPM (small sample size precluding results using the VCM).

**Table 4.** Comparison of the effectiveness^a^ of the monovalent XBB.1.5 COVID-19 vaccines against XBB and BA.2.86 VSLs among hospitalised SARI patients aged ≥65 years, by TSV, using the adapted predominance period (PP) with VSL category model (VCM) and with VSL proportion model (VPM) approaches, VEBIS hospital network, Europe

| Analysis model | TSV | VSL | OR | 95% CI | VE (%) | 95% CI | ROR (OR BA.2.86/OR XBB) | 95% CI | Median BA.2.86 (%)<sup>b</sup> | Median TSV (days) |
| --- | --- | --- | --- | --- | --- | --- | --- | --- | --- | --- |
| VCM | Overall: 14–59 days | BA.2.86 | 0.6 | 0.5–0.8 | 41 | 23–54 | 1.9 | 0.9–4.3 | 83 | 45 |
|  |  | XBB | 0.3 | 0.1–0.6 | 68 | 38–86 |  |  | 34 | 20 |
|  | 14–29 days | BA.2.86 | 0.6 | 0.4–1.1 | 36 | –8–62 | 2.6 | 1.0–7.6 | 77 | 23 |
|  |  | XBB | 0.2 | 0.1–0.6 | 75 | 45–90 |  |  | 34 | 19 |
|  | 30–59 days | BA.2.86 | 0.6 | 0.4–0.8 | 42 | 23–56 | NA<sup>c,d,e</sup> | NA<sup>c,d,e</sup> | 83 | 47 |
|  |  | XBB | NA<sup>c,d,e</sup> | NA<sup>c,d,e</sup> | NA<sup>c,d,e</sup> | NA<sup>c,d,e</sup> |  |  | 34 | 34 |
| VPM | Overall: 14–59 days | BA.2.86 | 0.5 | 0.4–0.7 | 48 | 35–59 | 1.8 | 1.1–2.9 | 70 | 45 |
|  |  | XBB | 0.3 | 0.2–0.5 | 71 | 50–83 |  |  | 30 | 21 |
|  | 14–29 days | BA.2.86 | 0.6 | 0.4–0.9 | 43 | 13–63 | 2.4 | 1.2–5.0 | 70 | 22 |
|  |  | XBB | 0.2 | 0.1–0.5 | 77 | 52–89 |  |  | 30 | 20 |
|  | 30–59 days | BA.2.86 | 0.5 | 0.4–0.7 | 46 | 30–59 | 1.4 | 0.7–2.7 | 70 | 47 |
|  |  | XBB | 0.4 | 0.2–0.8 | 62 | 16–83 |  |  | 30 | 32 |

CI: confidence interval; OR: odds ratio; PP: predominance period; ROR: ratio of odds ratios; TSV: Time since vaccination (at time of symptom onset); VE: vaccine effectiveness; VEBIS: Vaccine Effectiveness, Burden and Impact Studies; VSL: variant/sublineage
<sup>a</sup> The comparison of VE is measured by the ROR of vaccination between cases and controls against BA.2.86 and XBB VSLs ( $OR_{BA.2.86}/OR_{XBB}$ )
<sup>b</sup> Median proportion (%) of the BA.2.86 VSL during the period of analysis in each model approach
<sup>c</sup> Fewer than five vaccinated cases.
<sup>d</sup> Indication of small sample bias.
<sup>e</sup> <20 vaccinated patients.

The estimation of the ratio of ORs (*OR*_*BA*.2.86_/*OR_XBB_*) with the BA.2.86 variant proportion shows an increased risk of hospitalisation among those who were vaccinated and infected with BA.2.86 vs those vaccinated and infected with XBB VSLs, being statistically significant overall (14–59 days) and at 14–29 days post vaccination (Figure 3).

**Figure 3.**
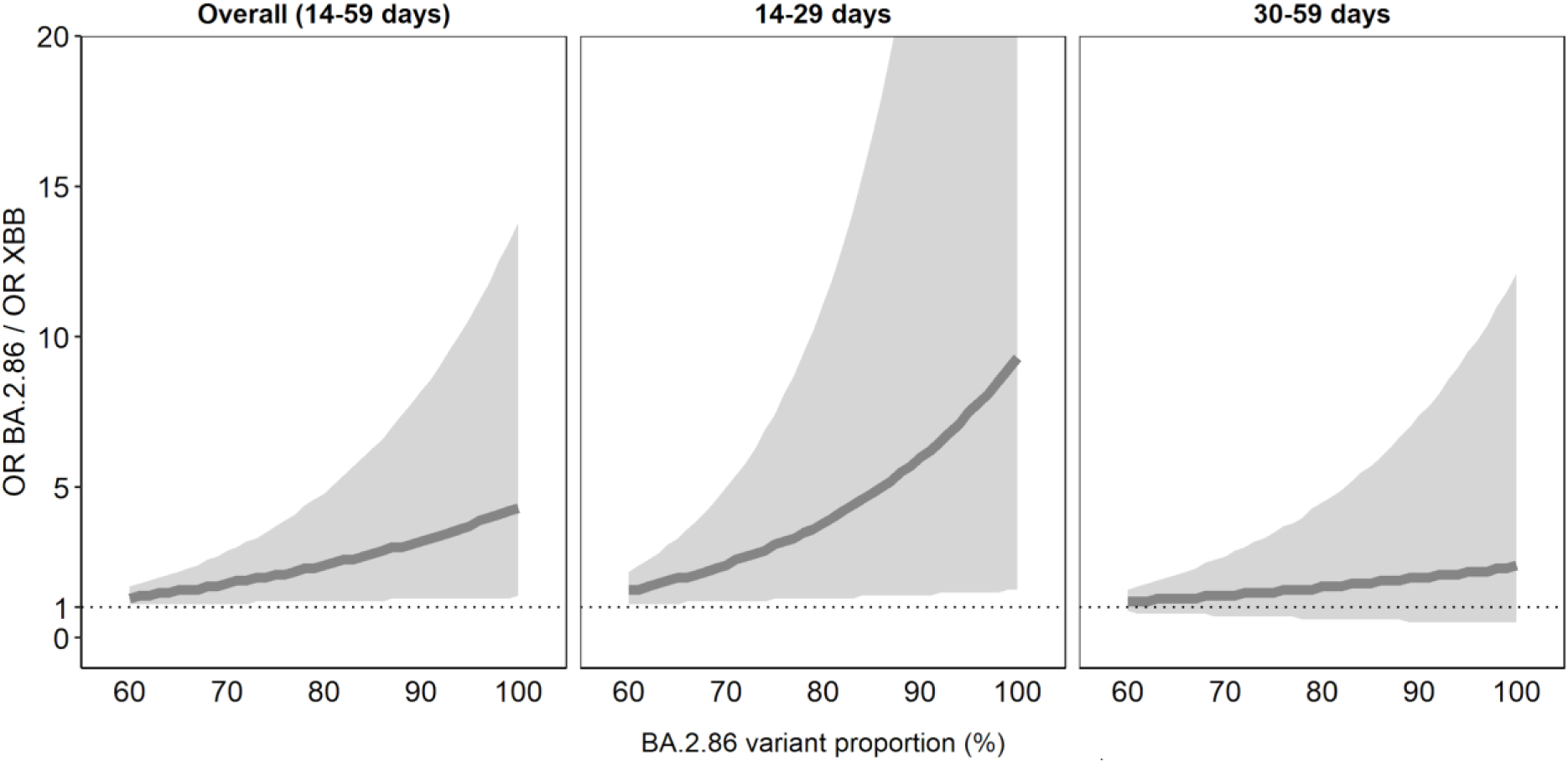
Comparison of the effectiveness^a^ of the monovalent XBB.1.5 COVID-19 vaccines against XBB and BA.2.86 VSLs among hospitalised SARI patients aged ≥65 years, by BA.2.86 VSLs using adapted PP with VPM^b^, by TSV, VEBIS hospital network, Europe OR: odds ratio; PP: standard predominance period; TSV: time since vaccination; VE: vaccine effectiveness; VEBIS: Vaccine Effectiveness, Burden and Impact Studies; VPM: VSL proportion model; VSL: variant/sublineage. ^a^ The comparison of the VE is measured by the ratio of the OR of vaccination between cases and controls against BA.2.86 and XBB VSLs (*OR*_*BA*.2.86_/*OR_XBB_* ), for the proportion of circulating BA.2.86 VSLs ≥60%. The ratio of OR is then calculated as *OR*_*BA*.2.86_/*OR_XBB_* = *OR_p_*/*OR*_1−*p*_, where *p* is the proportion of BA.2.86 VSLs. ^b^ The VPM model is a logistic regression model having case/control status as the dependent variable and the date of symptom onset, age, sex, presence of a chronic condition, vaccination status, VSL proportion and an interaction term between vaccination and the VSL proportion as independent variables. The form used to include date of symptom onset (linear, week, month, splines) and age (10-year age group, linear, splines) in the model was selected as the model with the lowest AIC.

## Discussion

We propose two different modelling approaches as an adaptation of the standard PP approach (VCM and the VPM), that allow for the estimation of VSL-specific VE as well as for statistical comparison of the VE against two different VSLs, when linked sequencing data are not available and inferring lineage through PCR techniques is not possible. We compared the VSL-specific VE results from our proposed models using the gold standard technique using linked SD and by using a proxy restricting the study period to periods of predominant circulation of the VSLs of interest (standard PP). Both validation techniques are well-established methodological approaches [1,4–11].

Results from our proposed modelling approaches produced similar VE estimates up to 59 days post vaccination (≤7% absolute difference) to those produced using established methods, varying from 68% to 71% for VE against XBB VSLs and from 41% to 48% for VE against BA.2.86 VSLs, using the VCM and with the VPM, respectively. Similar results from the different approaches were also found using narrower TSV intervals. Our study results also suggest that VE was lower against BA.2.86 than against XBB VSL hospitalisation, with estimates of the ROR of vaccination against BA.2.86 vs that against XBB ranging between 1.8 and 1.9 for the two proposed modelling approaches.

Other European studies and two US studies have found similar VSL comparative VE results; all suggested a lower VE against BA.2.86 than XBB VSLs [4,5,7,12], albeit with some differences in study design and population. Results from case-only studies also suggest lower VE against BA.2.86 than against XBB VSLs [17,18]. Huiberts *et al.* found vaccinated participants to have higher odds of BA.2.86 vs XBB infection, of 1.5 (95%CI: 0.8–2.6) [17], while Moustsen-Helms *et al.* estimated an OR of 1.5 (1.3–1.9) for BA.2.86 vs a non-BA.2.86 variant and 1.6 (1.3–2.0) for JN.1 lineage vs a non-BA.2.86 variant [18]. Estimates are slightly lower than ours; however, differences in study population, TSV, study period and study design (these studies report VE against infection, not hospitalisation) might all contribute to the differences found.

Both proposed models have many strengths: (1) they allow for the estimation of VE against each VSL in the same model, increasing the sample size used and consequently the precision, compared with the standard PP approach and VE using sequencing results (both of which use separate models for each study period). (2) The VPM, in our study, was able to provide VE estimates when standard PP and SD approaches were not able to do so due to low sample size. (3) The models also allow for a statistical comparison of VE against different VSLs, providing an estimate of a relative measure (the ratio of the OR of vaccination against two consecutively circulating VSLs), directly comparable with the OR reported from case–case studies. (4) Neither proposed model requires the use of linked sequencing and epidemiological data and the VPM does not require a threshold definition to determine the predominant VSL, which can be a limitation of the other techniques. (5) Finally, the VPM also allows the estimation of both indicators (VE and the ratio of the OR of vaccination against different VSLs) in a continuous form, providing more information about the measures of interest.

The proposed models also have limitations: (1) although the proposed methodology aims to overcome the lack of linked sequencing data to investigate VE differences against different VSLs, it still relies on SARS-CoV-2 sequencing data from surveillance made publicly available by countries, using a proxy for the VSL causing the infection at an ecologic level, which can introduce bias and may not be representative of the study population. (2) Using an external data source also requires having adequate sample size for robust measures of the VSL proportions. (3) These models are only useful with time-consecutive VSLs transitioning from predominance of one to the other. This method might not therefore be transferable to other pathogens not experiencing such transitions. For example, in seasonal influenza subclade VE estimation, subclades may be co-circulating rather than transitioning from dominance of one to another, rendering this method unfeasible. However, it would be interesting to test these models on VE against time-sequential transitioning seasonal influenza (sub)types, which can occur. It would also be useful to test these models on VE in other settings.

As in all observational studies, limitations also include the potential residual confounding that can remain even after adjustments (e.g. from vaccinated population differences over time). In addition, despite stratification by TSV in narrow 30-day time bands, adjustments for time of symptom onset and potential confounders, the inclusion of the underlying circulating VSL (included either as a categorical variable or as a proportion), and the short study period, it is unclear if we can fully disentangle the effect from TSV and the circulating VSL. This challenge is illustrated by the VPM results. We observe that precision of VE as a function of the BA.2.86 proportion decreases when the proportion of BA.2.86 increases at 14–29 days post-vaccination, and also decreases when the proportion of BA.2.86 is lower, at 30–59 days post-vaccination. However, for the overall analysis, the median TSV for the BA.2.86 VE estimate is higher than for the XBB estimate (45 vs 20–21 days), which could explain the differences in VE, despite the short overall period post vaccination. Using narrower TSV bands allows us to see a significant effect at 14–29 days, when median TSV is similar for both VSL estimates (varying between 19 and 23 days).

Further limitations were that we were unable to include all 12 sites in the analysis, and that there were different sites included in each of the validation approaches, which could lead to heterogeneity and lack of comparability of the analysis. However, the same sites were included for both VCM and VPM approaches, and results were similar between the proposed models and the standard techniques. In addition, the controls included in each approach may differ. For the SD, PP and VCM approaches, patient inclusion is performed by restricting patients over time, which means that some cases and controls recruited for each method may be different. In the SD approach, there might be some overlap of time and also of controls included for each of the XBB and BA.2.86 models. For both the standard PP and VCM approaches, however, controls do not overlap between VSL models by study site. while all cases and controls across both VSL periods are included in the VPM approach. Further analysis is required to investigate the impact of these changes in included sites and/or controls for each approach.

Small sample size precluded some results. In particular, it was not possible to estimate VE against XBB VSLs using SD due to no vaccinated cases, which would have been very relevant to validate our VE estimates against XBB. However, results from other studies have reported similar estimates [4], or report higher VE against XBB VSLs than against BA.2.86 (or JN.1) VSLs, despite differences in study design [5].

In conclusion, our two proposed novel modelling approaches provided similar VE estimates with generally improved precision compared with well-established methods. In particular, both VCM and VPM had greater precision than when using SD in our study, as we had fewer linked samples. The VPM was also able to provide VE estimates when both standard PP and SD techniques could not, due to low sample size. Although the optimal estimates would ideally integrate sequencing with epidemiological data, we anticipate that our novel modelling techniques will encourage other researchers lacking extensive study-specific sequencing data to provide more robust VE estimates.

## Supporting information

Section S1

## Data Availability

All data produced in the present study are available upon reasonable request to the authors.

## Acknowledgements

We thank all SARI patients and hospital teams included in these analyses. The Spanish team thanks all the participants in the SiVIRA Group (https://cne.isciii.es/documents/d/cne/colaboradores-sivira_2024-25-1) for Surveillance and Vaccine Effectiveness in Spain, including everyone involved in data collection and notification at the sentinel hospitals, laboratories, and public health units of all participating Autonomous Regions. We also extend our gratitude to the RELECOV network RELECOV (Red Española de Laboratorios de Secuenciación Genómica de SARS-CoV-2: https://relecov.isciii.es/es/) for their work in genomic sequencing and monitoring of SARS-CoV-2 in Spain. The Hungarian study team works as part of the National Laboratory for Health Security Hungary (RRF-2.3.1-21-2022-00006) supported by the National Research, Development and Innovation Office (NKFIH). The German team thanks all hospital teams who have contributed to German SARI surveillance, especially Torsten Bauer and David Krieger (Berlin Lung Institute, Respiratory Diseases Clinic Heckeshorn, Helios Klinikum Emil von Behring, Berlin). We want to thank the laboratory team at the National Reference Centre for Influenza and colleagues at the sequencing core facility of the Genome Competence Center, Robert Koch Institute (RKI), who contributed to the study. We sincerely appreciate the scientific support of Thomas Krannich, Marie Lataretu, Sofia Paraskevopoulou, and Dimitri Ternovoj from the Genome Competence Center, RKI, for their assistance with genome assembly. The Lithuanian team thanks Aistė Poškutė, Asta Stankauskaitė, Egidijus Balukevičius, Vilnius University Hospital Santaros Klinikos; Vilija Gurkšnienė, Vilnius University Faculty of Medicine, Vilnius. The Portuguese team thanks Camila Henriques, Débora Pereira, Margarida Tavares, Paula Pinto, and Cristina Bárbara. The Croatian team would like to express our gratitude to members of the National Influenza Laboratory and the Genomics and Bioinformatics Teams of the Microbiology Service, CIPH, who contributed to this study. The Romanian team thanks Mihaela Leuștean and Adina David, National Institute of Public Health Romania–National Public Health Laboratory, Bucharest.

## Ethical approval statement

The planning, conduct and reporting of the studies was in line with the Declaration of Helsinki. Official ethical approval was not required if studies were classified as being part of routine care/surveillance (Spain, Ireland, Malta); In Belgium and Germany, VE estimation is included in SARI surveillance. For Belgium, the study protocol was approved by the central Ethical Committee (CHU Saint-Pierre (AK/12-02-11/4111) initially in 2011, and UZ VUB (B.U.N. 143201215671) from 2014 on) and each participating hospital’s local ethical committees. The most recent amendment was approved on 27/9/2023 (reference 2012/310 Am6). The German SARI surveillance was approved by the Charité-Universitätsmedizin Berlin Ethical Board (Reference EA2/218/19). Other study sites obtained local ethical approval from a national review board (Croatia: 3 July 2023 by the Ethics committee of the Croatian Institute of Public Health, Class 030-02/23-01/3; Hungary: approved in March 2021 by the National Scientific and Ethical Committee for the period 01 September 2021–01 September 2024 (IV/1885-5/2021/EKU); Lithuania: approved 03 July 2020 by the Lithuanian Biomedical Research Ethics Committee No.: L-20-3/1-2; and later permission extended for the study period for seasons 2020–2024; Navarra: PI2020/45; Portugal: approved 19 January 2021 by the Ethics Committee of Instituto Nacional de Saúde Doutor Ricardo Jorge, no registration number given; Romania: approved by the Ethics Committee of the Ministerul Apărării Naionale Institutul Naional de Cercetare pentru Dezvoltare Medico-Militară, Cantacuzino” for the period 2022–2023, No. CE199/2022 and by the Ethics Committee of the National Institute of Public Health Romania No. 23468/2023).

## Contributing authors

**Belgium:** The BELSARI-Net group: Eva Bernaert, Reinout Naesens (Ziekenhuis Aan de Stroom, Antwerpen); Benedicte Lissoir, Catherine Sion, Sandra Koenig, Xavier Holemans (Grand Hôpital de Charleroi); Benedicte Delaere, Marc Bourgeois (CHU UCL Namur); Evelyn Petit, Marijke Reynders, Vanessa Verbeke (AZ Sint-Jan Brugge); Catherine Quoidbach, Francesco_Genderini, Gabriella Kollar, Nicolas Dauby, Stephanie Buylla (CHU St Pierre, Bruxelles), Sigi Van den Wijngaert (LHUB-ULB); Arne Witdouck, Deborah Degeyter, Eveline Vanhonacker, Lucie Seyler, Siel Daelemans (UZ Brussel); Marieke Bleyen , Marlies Blommen, Natasja Detillieu, Veerle Penders (Jessa Ziekenhuis Hasselt); Dylan Lievens, Marie-Pierre Parsy, Melanie Delvallee, Pierre Struyven (CHWAPI, Tournai); Isabel Leroux-Roels, Pascal De Waegemaeker, Silke Ternest (UZ Gent); Anna Parys, Francois Dufrasne, Sarah Denayer, Claire Brugerolles, Laurane De Mot, Mathil Vandromme, Sebastien Fierens, Sven Hanoteaux, Yves Lafort, Nathalie Bossuyt (Sciensano). **Croatia:** Maja Ilić, Ivan Mlinarić, Dragan Jurić, Irena Tabain (Croatian Institute of Public Health); Diana Nonković, Petra Tomaš Petrić (Teaching Public Health Institute of Split-Dalmatia County); Suzana ladinov, Matea Nikolić, Ana Brnas, Svjetlana Karabuva, Mihaela Čikeš, Marija Tonkić (University Hospital of Split). **Epiconcept, France:** E Kissling, J Howard, A Nardone. **Germany:** Silke Buda, Kristin Tolksdorf, Annika Erdwiens, Ute Preuss (Department for Infectious Disease Epidemiology, RKI); Djin-Ye Oh, Janine Reiche (National Reference Centre for Influenza, Robert Koch Institute). **Hungary:** Viktória Velkey, Katalin Kristóf, Bánk Fenyves, Csaba Varga, Krisztina Mucsányiné Juhász, Csaba Luca, Katalin Krisztalovics (Semmelweis University). **Lithuania:** Aukse Mickiene, Roberta Vaikutyte-Ramanauskiene (Department of Infectious Diseases, Lithuanian University of Health Sciences, Kaunas, Lithuania); Fausta Majauskaite (Infectious Diseases and Dermatovenerology, Institute of Clinical Medicine, Medical Faculty, Vilnius University, Vilnius)**. Malta:** Tanya Melillo, Ariana Wijermans, John Paul Cauchi, Stephen Abela, Gerd Xuereb. **Navarre region, Spain:** Aitziber Echeverría, Camino Trobajo-Sanmartín, Nerea Egüés, Alba Gasque-Satrústegui, Naroa Andueza, Guillermo Ezpeleta (Instituto de Salud Pública de Navarra – IdiSNA – CIBERESP); Ana Navascués, Carmen Ezpeleta (Hospital Universitario de Navarra – IdiSNA). **Portugal:** Ana Paula Rodrigues (Department of Epidemiology); Nuno Verdasca, Victor Borges (Infectious Diseases Department). **Romania:** Isabela Ioana Loghin (“Saint Parascheva” Clinical Hospital for Infectious Diseases, Iasi); Corneliu Popescu, Gratiela Tardei, Marin Alexandru (“Dr Victor Babes” Clinical Hospital of Infectious and Tropical Diseases, Bucharest); Sorin Dinu, Catalina Pascu, Alina Ivanciuc, Carmen Maria Cherciu, Iulia Bistriceanu, Mihaela Oprea, Maria Elena Mihai (“Cantacuzino” National Military–Medical Institute for Research and Development, Bucharest). **Spain:** Susana Monge, Gloria Pérez-Gimeno, Marcos Lozano, Nuria Rincón Calvo, Mª del Carmen Pacheco Martínez.

