## Supplementary Table S3 for "Modelling approaches for estimating vaccine effectiveness of consecutive SARS-CoV-2 variant sublineages in the absence of study-specific genetic sequencing data, VEBIS hospital network, Europe, 2023/24"

Supplementary Table S3. Start date[a] of the XBB and BA.2.86 VSL predominant periods for a 60% predominance threshold,[b] by site, VEBIS hospital study, Europe

| Site | BA.2.86 First date in start week | XBB First date in start week | XBB Last date in end week |
|---|---|---|---|
| Belgium (BE) | 27 Nov 2023 | 27 Feb 2023 | 12 Nov 2023 |
| Czechia (CZ) | 11 Dec 2023 | 06 Feb 2023 | 26 Nov 2023 |
| Germany (DE) | 04 Dec 2023 | 06 Mar 2023 | 26 Nov 2023 |
| Spain (ES) | 04 Dec 2023 | 27 Feb 2023 | 12 Nov 2023 |
| Croatia (HR) | 01 Jan 2024[c] | 27 Feb 2023 | 10 Dec 2023 |
| Hungary (HU) | 25 Dec 2023[c] | 27 Mar 2023[c] | 10 Dec 2023[c] |
| Ireland (IE) | 11 Dec 2023 | 13 Feb 2023 | 26 Nov 2023 |
| Lithuania (LT) | 25 Dec 2023 | 20 Mar 2023 | 03 Dec 2023 |
| Malta (MT) | 25 Dec 2023[c] | 13 Mar 2023[c] | 03 Dec 2023[c] |
| Portugal (PT) | 27 Nov 2023[c] | 13 Mar 2023 | 15 Oct 2023 |
| Romania (RO) | 01 Jan 2024[c] | 06 Mar 2023[c] | 17 Dec 2023[c] |

AT: Austria; BG: Bulgaria; EL: Greece; ES: Spain; HR: Croatia; HU: Hungary; IT: Italy; MT: Malta; PT: Portugal; RO: Romania; SI: Slovenia; SK: Slovakia; VEBIS: Vaccine Effectiveness, Burden and Impact Studies; VSL: variant/sublineage.

[a] Date of first day of the start week.

[b] Data on SARS-CoV-2 circulation available from ECDC ERVISS Github, extracted on 16 May 2024.

[c] Start date week was calculated based on data from the site country and its neighbouring countries together: HR (neighbours = HU, SI); HU (neighbours = AT, HR, RO, SI, SK); MT (neighbours = HR, EL, IT); PT (neighbours = ES); RO (neighbours = BG, HU).
