## Supplementary Table S4 for "Modelling approaches for estimating vaccine effectiveness of consecutive SARS-CoV-2 variant sublineages in the absence of study-specific genetic sequencing data, VEBIS hospital network, Europe, 2023/24"

### Demonstration of ratio of odds ratios (ORs) in case–case studies

**Supplementary Table S4.** Example of a table with the number of XBB and BA.2.86 cases and controls by their vaccination status, VEBIS hospital study, Europe

| Vaccination status | XBB cases | BA.2.86 cases | Controls |
|---|---|---|---|
| Vaccinated | a | b | c |
| Unvaccinated | d | e | f |

In case–case studies, the odds of vaccination between BA.2.86 and XBB VSLs ($OR_{BA.2.86\quad vs\quad XBB}$) is given by Eq. S2:

$$OR_{BA.2.86\quad vs\quad XBB} = \frac{bd}{ea} \quad (\mathrm{Eq.\,S2})$$

The ratio of the OR between the two variants ($\frac{OR_{BA.2.86}}{OR_{XBB}}$) is given by Eq. S3, which amounts to the same in Eq. S2:

$$\frac{OR_{BA.2.86}}{OR_{XBB}} = \frac{\frac{bf}{ce}}{\frac{af}{cd}} = \frac{bd}{ea} \quad (\mathrm{Eq.\,S3})$$

where the OR of vaccination against XBB ($OR_{XBB}$) and BA.2.86 ($OR_{BA.2.86}$) VSLs are given by:

$$OR_{XBB} = \frac{af}{cd} \quad (\mathrm{Eq.\,S4})$$

$$OR_{BA.2.86} = \frac{bf}{ce} \quad (\mathrm{Eq.\,S5})$$
