## Supplementary Figure S1 for "Modelling approaches for estimating vaccine effectiveness of consecutive SARS-CoV-2 variant sublineages in the absence of study-specific genetic sequencing data, VEBIS hospital network, Europe, 2023/24"

**Supplementary Figure S1.** Weekly BA.2.86 VSL proportion reported by the country, the proportion calculated using neighbouring countries and the final estimated time series using logistic regression used in the category and proportion models, based on TESSy/GISAID data (ECDC ERVISS Github), VEBIS hospital network, Europe.

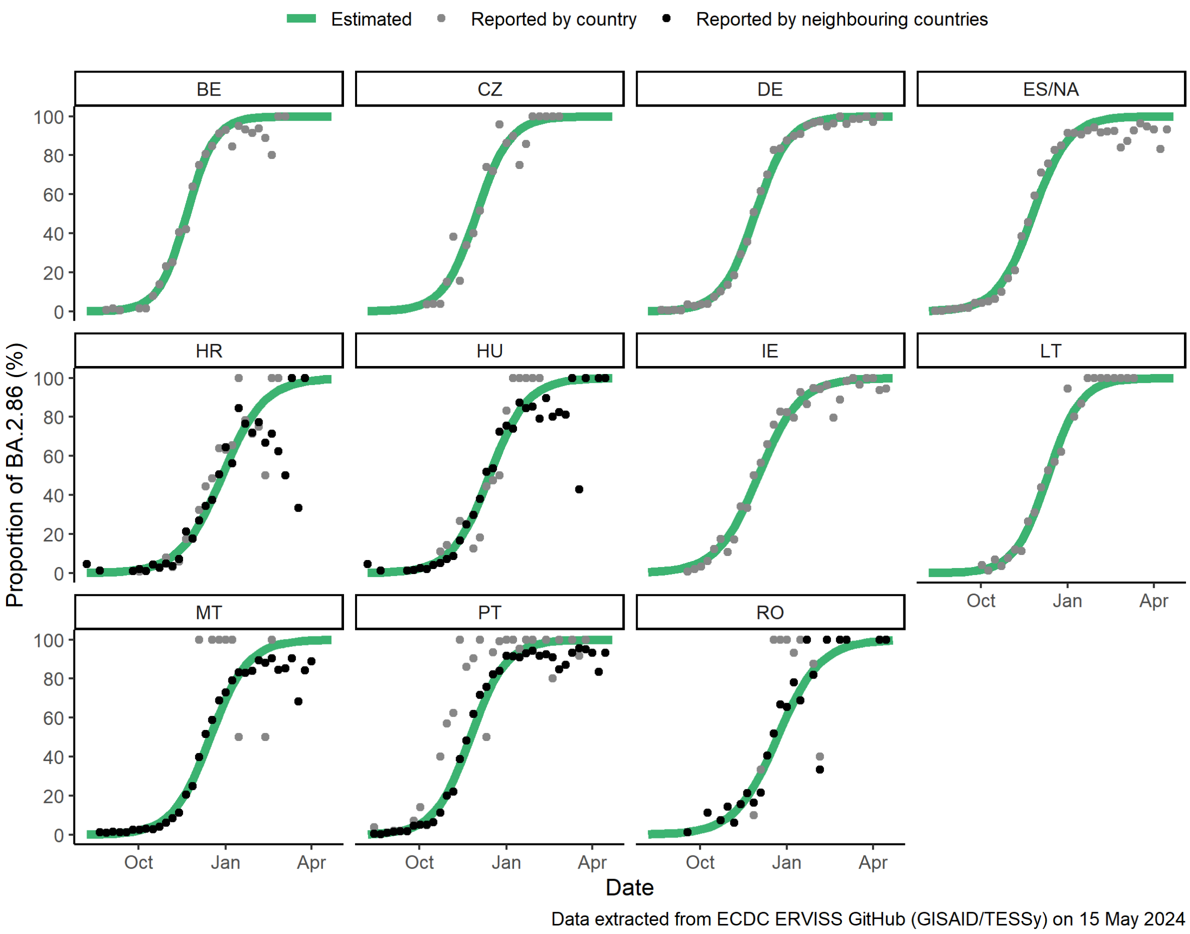


The variant proportion data was sourced from ERVISS Github [1].

The model equation used to estimate the weekly time series for each country is presented in Eq. S1, where p is the reported proportion of BA.2.86 (either from the country or, when missing data/low number of sequenced samples, based on the country and its neighbours together), and t represents time in weeks.

$$\log\left(\frac{p}{1-p}\right) = \beta_0 + \beta_1 t \quad (\text{Eq. S1})$$
