## Supplementary Figure S4 for "Modelling approaches for estimating vaccine effectiveness of consecutive SARS-CoV-2 variant sublineages in the absence of study-specific genetic sequencing data, VEBIS hospital network, Europe, 2023/24"

Supplementary Figure S4. Patient exclusion flowchart for the XBB standard predominance period (standard PP) analysis, VEBIS hospital study, Europe.

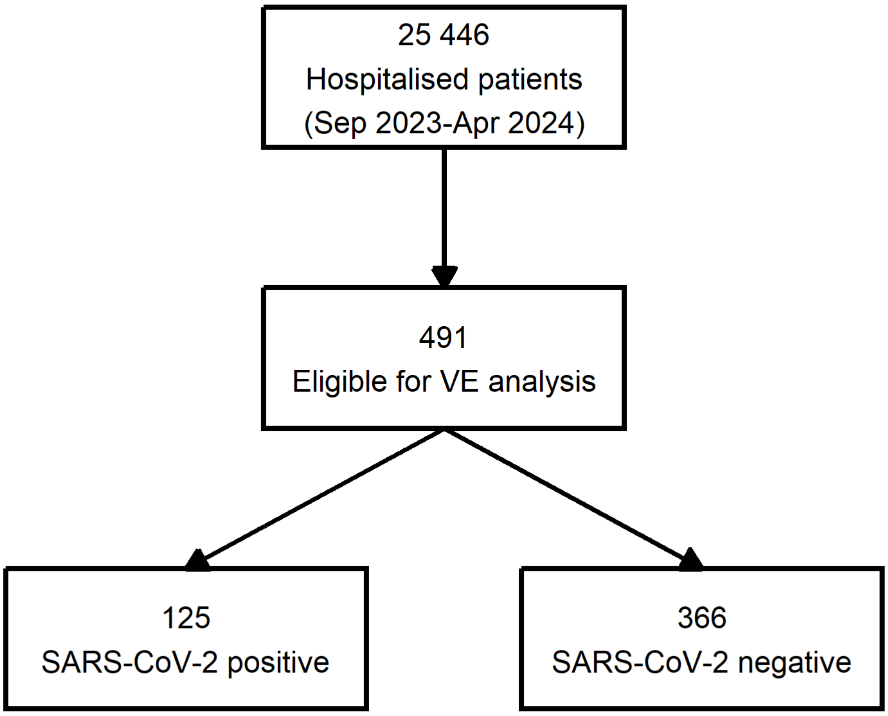


**24 955 patients excluded**

**23 467 outside the analysis period**
(1416 with date of symptom onset before the start of the campaign + 14 days, 405 from 1 site with vaccination data issues before 13 November 2023 and 21646 XBB predominant period)

**310 exclusion criteria (protocol)**
(94 missing information on SARI case definition, 152 did not meet the ECDC SARI case definition and 64 had a missing RT-PCR test result/did not have an RT-PCR test)

**741 in an ineligible population group**
(5 healthcare workers, 75 residents in a long-term care facility and 661 aged <65 years)

**192 missing key covariates for analysis**
(2 missing admission date, 18 missing symptom onset date, 2 missing age, 2 missing sex, 25 missing information on common chronic diseases, 25 missing vaccination status and 118 missing date of last received vaccine dose)

**55 were ineligible due to timing of symptom onset, swab and hospitalisation**
(28 swabbed >10 days after symptom onset, 3 swabbed >3 days before symptom onset, 23 swabbed >48h after hospitalisation and 1 swabbed >14 days before hospitalisation)

**90 with ineligible vaccination status**
(75 with last vaccine dose <14 days before symptom onset and 15 with last vaccine dose received within 180 prior to the campaign)

**100 due to site restrictions**
(5 sites with fewer than 5 cases or controls (n=86) and 1 site with no vaccinated cases and controls (n=14))

**Records included are from 41 hospitals in four sites (Germany, Spain, Croatia, Navarre region and Spain)**
