## Supplementary Figure S5 for "Modelling approaches for estimating vaccine effectiveness of consecutive SARS-CoV-2 variant sublineages in the absence of study-specific genetic sequencing data, VEBIS hospital network, Europe, 2023/24"

Supplementary Figure S5. Patient exclusion flowchart for the BA.2.86 standard predominance period (standard PP) analysis, VEBIS hospital study, Europe.

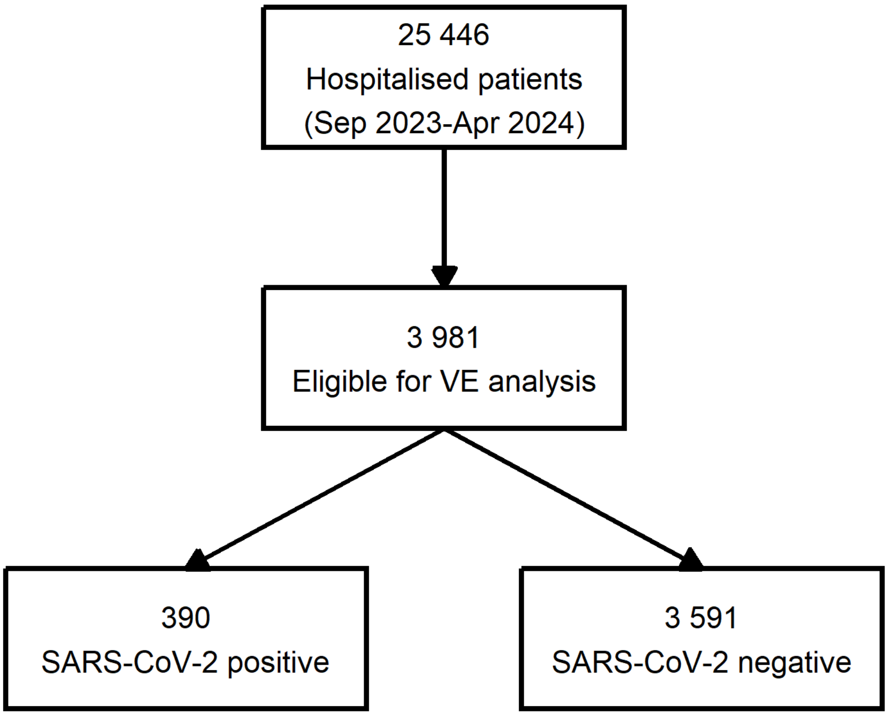


**21 465 patients excluded**

**6 103 outside the analysis period**
(1416 with date of symptom onset before the start of the campaign + 14 days, 405 from 1 site with vaccination data issues before 13 November 2023 and 4282 BA.2.86 predominant period)

**2 616 exclusion criteria (protocol)**
(700 missing information on SARI case definition, 951 did not meet the ECDC SARI case definition and 965 had a missing RT-PCR test result/did not have an RT-PCR test)

**7 388 in an ineligible population group**
(38 healthcare workers, 737 residents in a long-term care facility and 6613 aged <65 years)

**929 missing key covariates for analysis**
(11 missing admission date, 106 missing symptom onset date, 13 missing age, 7 missing sex, 270 missing information on common chronic diseases, 239 missing vaccination status and 283 missing date of last received vaccine dose)

**374 were ineligible due to timing of symptom onset, swab and hospitalisation**
(260 swabbed >10 days after symptom onset, 19 swabbed >3 days before symptom onset and 95 swabbed >48h after hospitalisation)

**3 496 with ineligible vaccination status**
(58 with last vaccine dose <14 days before symptom onset, 1 with contraindications for vaccination, 47 with last vaccine dose received within 180 days prior to the campaign, 9 not eligible to be vaccinated in PT and IE, 2 not vaccinated with a booster dose during the XBB.1.5 vaccination campaign in PT and IE, 18 vaccinated with bivalent vaccine, 1 vaccinated with vaccine brand other than Comirnaty, Spikevax or Nuvaxovid, 1 received Comirnaty XBB.1.5 vaccine before 31 August 2023 and 3359 vaccinated ≥ 60 days before onset)

**559 due to site restrictions**

(1 site with fewer than 5 cases or controls (n=166) and 5 sites with no vaccinated cases and controls (n=393))

| |
|---|
| **21 465 patients excluded** |
| **Records included are from 60 hospitals in six sites (Belgium, Germany, Spain, Ireland, Malta, Navarre region and Spain)** |
