## Supplementary Figure S6 for "Modelling approaches for estimating vaccine effectiveness of consecutive SARS-CoV-2 variant sublineages in the absence of study-specific genetic sequencing data, VEBIS hospital network, Europe, 2023/24"

Supplementary Figure S6. Patient exclusion flowchart for the XBB and BA.2.86 adapted predominance period (adapted PP) variant/sublineage category model (VCM) analysis, VEBIS hospital study, Europe.

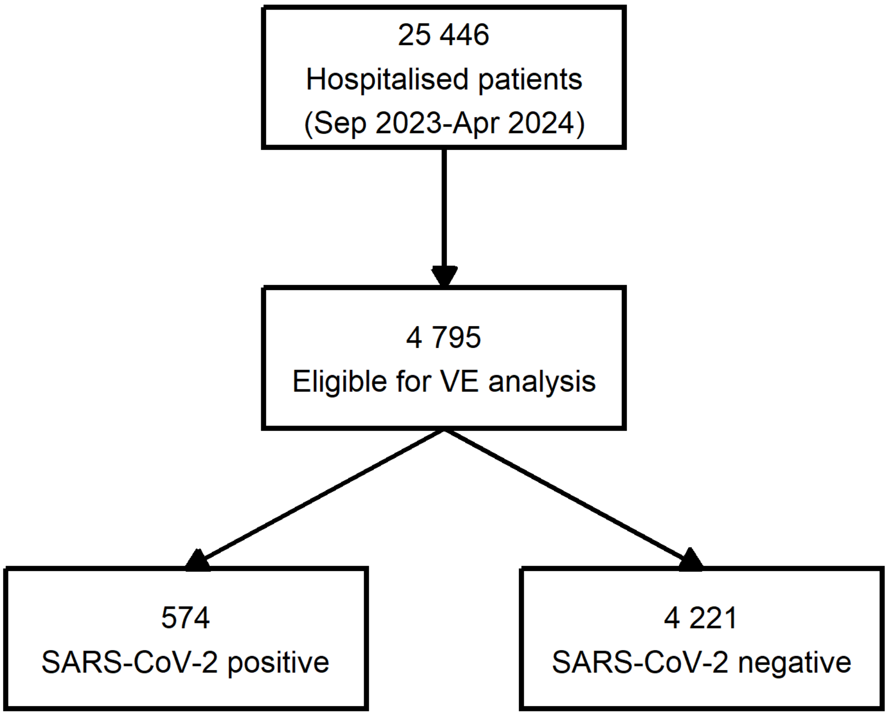


**20 651 patients excluded**

**3 477 outside the analysis period**

(1 416 with date of symptom onset before the start of the campaign + 14 days, 405 from 1 site with vaccination data issues before 13 November 2023 and 1 656 between 41% and 59% BA.2.86 circulating)

**3 025 exclusion criteria (protocol)**

(808 missing information on SARI case definition, 1 145 did not meet the ECDC SARI case definition and 1 072 had a missing RT-PCR test result/did not have an RT-PCR test)

**8 402 in an ineligible population group**

(43 healthcare workers, 853 residents in a long-term care facility and 7 506 aged <65 years)

**1 137 missing key covariates for analysis**

(13 missing admission date, 122 missing symptom onset date, 15 missing age, 8 missing sex, 299 missing information on common chronic diseases, 266 missing vaccination status and 414 missing date of last received vaccine dose)

**445 were ineligible due to timing of symptom onset, swab and hospitalisation**

(299 swabbed >10 days after symptom onset, 23 swabbed >3 days before symptom onset, 122 swabbed >48 hrs after hospitalisation and 1 swabbed >14 days before hospitalisation)

**3 614 with ineligible vaccination status**

(158 with last vaccine dose <14 days before symptom onset, 1 with contraindications for vaccination, 63 with last vaccine dose received within 180 days prior to the campaign, 11 not eligible to be vaccinated in PT and IE, 2 not vaccinated with a booster dose during the XBB.1.5 vaccination campaign in PT and IE, 18 vaccinated with bivalent vaccine, 1 vaccinated with vaccine brand other than Comirnaty, Spikevax or Nuvaxovid, 1 received Comirnaty XBB.1.5 vaccine before 31 August 2023 and 3 359 vaccinated ≥ 60 days before onset)

**551 due to site restrictions**

| |
|---|
| **20 651 patients excluded** |
| (1 site with fewer than 5 cases or controls (n=198) and 4 sites with no vaccinated cases and controls (n=353)) |

**Records included are from 62 hospitals in seven sites (Belgium, Germany, Spain, Croatia, Ireland, Malta, Navarre region and Spain)**
