## Supplementary Figure S7 for "Modelling approaches for estimating vaccine effectiveness of consecutive SARS-CoV-2 variant sublineages in the absence of study-specific genetic sequencing data, VEBIS hospital network, Europe, 2023/24"

Supplementary Figure S7. Patient exclusion flowchart for the XBB and BA.2.86 adapted predominance period (adapted PP) variant/sublineage proportion model (VPM) analysis, VEBIS hospital study, Europe.

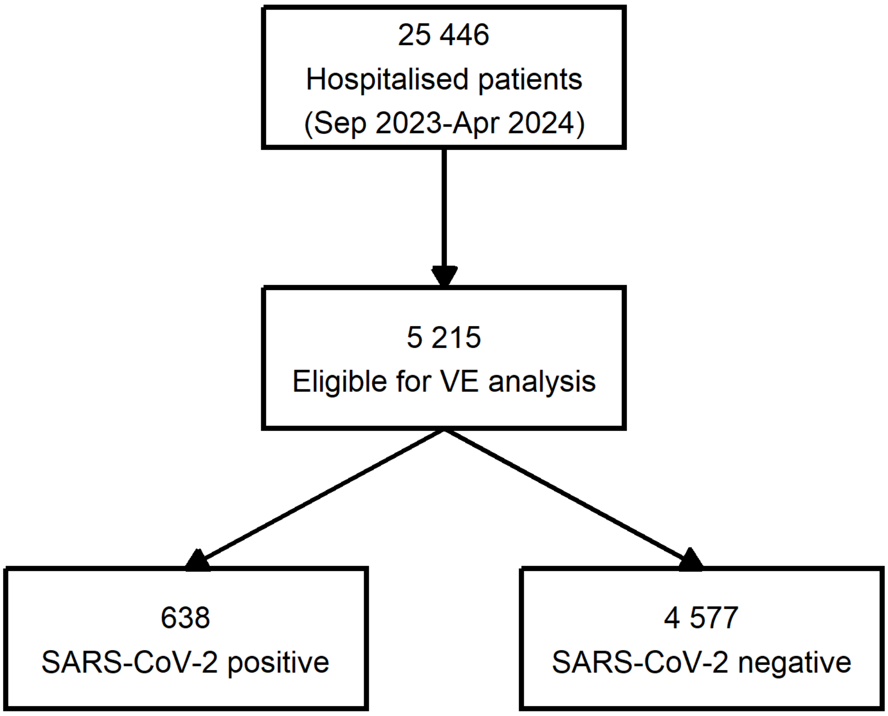


**20 231 patients excluded**

**1 821 outside the analysis period**
(1 416 with date of symptom onset before the start of the campaign + 14 days and 405 from 1 site with vaccination data issues before 13 November 2023)

**3 436 exclusion criteria (protocol)**
(19 missing consent information, 855 missing information on SARI case definition, 1231 did not meet the ECDC SARI case definition and 1 331 had a missing RT-PCR test result/did not have an RT-PCR test)

**9 045 in an ineligible population group**
(46 healthcare workers, 925 residents in a long-term care facility and 8 074 aged <65 years)

**1 199 missing key covariates for analysis**
(3 missing swab date, 13 missing admission date, 127 missing symptom onset date, 16 missing age, 9 missing sex, 306 missing information on common chronic diseases, 287 missing vaccination status and 438 missing date of last received vaccine dose)

**471 were ineligible due to timing of symptom onset, swab and hospitalisation**
(316 swabbed >10 days after symptom onset, 24 swabbed >3 days before symptom onset, 130 swabbed >48 hrs after hospitalisation and 1 swabbed >14 days before hospitalisation)

**3 663 with ineligible vaccination status**
(193 with last vaccine dose <14 days before symptom onset, 1 with contraindications for vaccination, 69 with last vaccine dose received within 180 days prior to the campaign, 13 not eligible to be vaccinated in PT and IE, 2 not vaccinated with a booster dose during the XBB.1.5 vaccination campaign in PT and IE, 20 vaccinated with bivalent vaccine, 1 vaccinated with vaccine brand other than Comirnaty, Spikevax or Nuvaxovid, 1 received Comirnaty XBB.1.5 vaccine before 31 August 2023 and 3 363 vaccinated ≥ 60 days before onset)

**596 due to site restrictions**

| **20 231 patients excluded** |
| --- |
| (1 site with fewer than 5 cases or controls (n=232) and 4 sites with no vaccinated cases and controls (n=364)) |
| **Records included are from 63 hospitals in seven sites (Belgium, Germany, Spain, Croatia, Ireland, Malta, Navarre region and Spain)** |
