## Supplementary material for "Modelling approaches for estimating vaccine effectiveness of consecutive SARS-CoV-2 variant sublineages in the absence of study-specific genetic sequencing data, VEBIS hospital network, Europe, 2023/24": Section S1

**Supplementary Section S1.** Description of the proxy determination for XBB and BA.2.86 predominance periods

The study periods were restricted to the weeks where the consecutive variant sublineages (VSLs) of interest were predominant. We defined weekly thresholds for both XBB and BA.2.86 VSL predominant periods as ≥ 60%, by country (Figure S1). We excluded weeks with a low number of sequenced samples (n<17) to determine the start/end of the predominance period and, for countries with low quality data, used data from neighbouring countries, in addition to their own data, to produce the final variant proportion time series (Figure S1).
