## Supplementary Table S1 for "Modelling approaches for estimating vaccine effectiveness of consecutive SARS-CoV-2 variant sublineages in the absence of study-specific genetic sequencing data, VEBIS hospital network, Europe, 2023/24"

**Supplementary Table S1.** Definitions used for analyses, including analytic exclusions, VEBIS hospital study, Europe

| | Definition |
|---|---|
| SARI patients | Patients hospitalised for ≥24 hours with at least one of the following symptoms: fever, cough, shortness of breath, or sudden onset of anosmia, ageusia, or dysgeusia[a] |
| Case | SARI patients testing positive for SARS-CoV-2 by RT-PCR within 48 hours of admission or in the previous 14 days |
| Control | SARI patients PCR-negative for SARS-CoV-2 by RT-PCR within 48 hours of admission with no positive test in the previous 14 days |
| Vaccinated | Last COVID-19 vaccine dose received after the introduction of the XBB.1.5 vaccine in patient's country, ≥14 days before symptom onset |
| Unvaccinated | Never received a COVID-19 vaccine (with the exception of IE and PT[b]), or last COVID-19 vaccine dose received ≥180 days before the start of the vaccination campaign in each country[c,d] |
| Any chronic condition | Patients with at least one condition out of diabetes, heart disease, lung disease/asthma, or who are immunocompromised |
| No chronic condition | None of the conditions listed above |
| Excluded based on vaccination status | • Patients vaccinated 1–13 days before symptom onset<br>• Where vaccine product and type was known, those vaccinated with vaccine other than adapted XBB.1.5 during the autumn 2023 vaccination campaign<br>• Where only vaccine product was known, those vaccinated with brands other than Comirnaty, Spikevax and Nuvaxovid during the autumn 2023 vaccination campaign<br>• Patients with fewer than two doses of a COVID-19 vaccine in IE and PT[b] |
| Excluded for missing/ erroneous key variables | • Missing/erroneous information on variables included in the analysis (sex, age, chronic conditions, and dates of onset, swab and hospital admission)<br>• Missing/erroneous information on variables used to determine eligible vaccination (vaccination status or date) |
| Excluded for other reasons | • Sites with fewer than five cases or controls<br>• Sites with no vaccinated SARI patients in both case and control groups[d] |

IE: Ireland; PT: Portugal; SARI: severe acute respiratory infection; VEBIS: Vaccine Effectiveness, Burden and Impact Studies.

[a] Peralta-Santos A. Assessment of COVID-19 surveillance case definitions and data reporting in the European Union. Briefing requested by the ENVI committee. Brussels: European Parliament; July 2020. http://www.europarl.europa.eu/RegData/etudes/BRIE/2020/652725/IPOL_BRI(2020)652725_EN.pdf

[b] In these countries, XBB.1.5 vaccine was only available to those who had at least completed the primary vaccination course for COVID-19, so patients who received fewer than two doses of a COVID-19 vaccine from sites in IE and PT were excluded.

[c] For vaccination campaign start dates by country/site, see Supplementary material Table S2.

[d] For inclusion flowcharts for each analysis, see Supplementary material Figures S1–S5.
