## Supplementary Table S2 for "Modelling approaches for estimating vaccine effectiveness of consecutive SARS-CoV-2 variant sublineages in the absence of study-specific genetic sequencing data, VEBIS hospital network, Europe, 2023/24"

Supplementary Table S2. Start date of the 2023/24 vaccination campaign by site, VEBIS hospital study, Europe.

| Site | Date vaccination campaign started | XBB.1.5 vaccine introduction |
|---|---|---|
| Belgium (BE) | 15 Sep 2023 | 15 Sep 2023 |
| Czechia (CZ) | 04 Aug 2023 | 04 Aug 2023 |
| Germany (DE) | 18 Sep 2023 | 18 Sep 2023 |
| Spain (ES) | 25 Sep 2023 | 25 Sep 2023 |
| Croatia (HR) | 18 Sep 2023 | 18 Sep 2023 |
| Hungary (HU) | 01 Oct 2023 | 01 Dec 2023 |
| Ireland (IE) | 02 Oct 2023 | 02 Oct 2023 |
| Lithuania (LT) | 05 Oct 2023 | 05 Oct 2023 |
| Malta (MT) | 09 Oct 2023 | 09 Oct 2023 |
| Navarre region, Spain (NA)[a] | 16 Oct 2023 | 16 Oct 2023 |
| Portugal (PT) | 29 Sep 2023 | 29 Sep 2023 |
| Romania (RO) | 02 Oct 2023 | 17 Jan 2024 |

[a] The 2023 autumn vaccination campaign started earlier in the Navarre region in Spain (25 September 2023), with sparse administration of vaccines. Vaccines started being administered in larger quantities after 16 October 2023.
